# A randomised controlled trial of interventions to promote adoption of physical activity in adults with severe mental illness

**DOI:** 10.1101/2024.07.25.24310981

**Authors:** Justin J. Chapman, Aaron Miatke, Dorothea Dumuid, Jairo Migueles, Shuichi Suetani, Nicole Korman, Mike Trott, Jacqueline Byrne, Dan Siskind, Donni Johnston, Jeanette Sewell, Michael Breakspear, Sue Patterson

## Abstract

**Background and aims:** Adults with severe mental illness (SMI) have lower physical activity (PA) than the general population. Supervised exercise interventions provide high support but may not effectively promote motivation, which is important for behaviour change. Motivational strategies such as PA counselling may target motivation more directly; however, the effectiveness in people with SMI is unclear.

**Methods:** This was a randomised controlled trial of interventions designed to promote PA in adults with SMI. Participants were randomised to either: (1) supervised exercise (GYM), or (2) motivational counselling and self-monitoring using fitness trackers (MOT). Group sessions were once/week over 8-weeks. The primary outcome was time spent in moderate-to-vigorous PA (MVPA) assessed using GENEActiv accelerometers worn continuously. Change in MVPA was assessed using the cumulative change from baseline, and as a composition of light activity, sedentary behaviour, and sleep.

**Results:** Sixty-four participants were allocated (63% male, 82% overweight/obese, 59% psychotic disorder). Accelerometer-derived MVPA increased for the MOT group between baseline and post-intervention, and the cumulative sum of change in MVPA from baseline in the MOT group was higher than the GYM group. Compositional analyses showed stable weekly activity profiles, with no significant changes attributable to group allocation.

**Conclusions:** The cumulative change in MVPA was higher for MOT than GYM; however, compositional analyses that considers MVPA as a composition of other daily behaviours showed no change in composition over the intervention period. Exercise interventions should incorporate motivational strategies and supervised exercise; future research should investigate behaviour change interventions with longer durations and more frequent sessions.

**Registration details:** The trial is registered under the Australian and New Zealand Clinical Trial Registry (ACTRN12617001017314).

## INTRODUCTION

Severe mental illness (SMI) is characterised by significant functional impairment (Ruggeri et al., 2000), and typical diagnoses including psychotic disorders, bipolar disorder, and major depressive disorder. People with SMI experience a 10-20 year shorter life expectancy than the general population which is largely caused by preventable cardiometabolic conditions (Walker et al., 2015): people with SMI are at 2-3 times higher risk of developing diabetes and cardiovascular disease compared with the general population (Firth et al., 2019). Physical activity (PA) and exercise (structured PA to enhance fitness) can help prevent and manage cardiometabolic conditions (Lavie et al., 2019). Supervised exercise interventions, in which a health professional provides instruction for exercise, can support people to exercise (Alexandratos et al., 2012) and are feasible for people with SMI, with reported dropout rates of exercise trials being comparable to psychological therapies (Cella et al., 2023) and medication trials (Leucht et al., 2012). However, exercise professionals are not routinely employed in public mental health services, and accessing professional exercise support in the community may be difficult for people with SMI (Firth et al., 2016). Further, it is unclear how supervised exercise influences behaviour change outside supervised sessions and once supervised support ceases. PA counselling may be a more accessible form of exercise support that addresses motivation for PA, which may support longer term behaviour change (Vancampfort et al., 2015).

Lifestyle counselling or coaching programs utilise motivational approaches to enhance self-awareness and capabilities to make positive changes, and are commonly founded on goal-setting theory and other behaviour change frameworks (Frates et al., 2011). There are numerous behaviour change techniques (BCTs) that can be utilised (Michie, Ashford, et al., 2011), and evidence suggests that interventions involving self-monitoring combined with other self-regulatory techniques (e.g., goal setting) may be more effective than interventions without these techniques (Greaves et al., 2011; Michie et al., 2009). PA counselling programs may be more easily implemented in routine mental health care because of lower need for dedicated space or equipment, and can be delivered by a wider variety of health professionals (Lee et al., 2014; Temmingh et al., 2013). Reviews of PA counselling have demonstrated improved health outcomes in people with chronic health conditions (O’Halloran et al., 2014), and preliminary evidence indicates potential for increasing PA in people SMI (Ashdown-Franks et al., 2018). While PA counselling may be less resource intensive than supervised exercise, it is unclear whether counselling approaches are as effective as supervised exercise for facilitating behaviour change in people with SMI. Comparing supervised exercise and counselling approaches could inform the design of PA interventions for this group.

The primary aim of this study was to compare the efficacy of two interventions to promote PA in people with SMI: a supervised exercise intervention, and a motivational PA counselling intervention. The primary outcome was accelerometer-derived moderate-to-vigorous physical activity (MVPA). A secondary aim was to compare the impact of the interventions on motivation for PA.

## METHODS AND ANALYSIS

### Study design

This was a pre-registered two-arm, parallel group, randomised controlled trial (RCT) of interventions designed to promote adoption and maintenance of PA among adults with SMI. Ethical approval was granted by the Royal Brisbane Women’s Hospital Human Research Ethics Committee (HREC/17/QRBW/302); the trial was registered under the Australian and New Zealand Clinical Trial Registry (ACTRN12617001017314).

### Setting and participants

Participants were recruited from public mental health service teams (Metro North Mental Health, and Metro South Addictions and Mental Health in Brisbane, Australia) which provide outpatient care for people with SMI. The study was promoted at clinical team meetings, and staff were asked to refer potentially eligible individuals from October 2017 to January 2020. Inclusion criteria were aged 18-65 years and sufficiently fluent in English to provide informed consent. Exclusion criteria were: i) receiving treatment for an eating disorder (because exercise can be contraindicated for some eating disorders); and ii) exceeding physical activity guidelines of more than 300 minutes of self-reported MVPA in the previous week. Screening involved assessment of PA over the phone using an adapted version of the Active Australia questionnaire (Australian Institute of Health and Welfare, 2004), which asked about time spent in walking, moderate and vigorous activities in the previous week.

Eligible individuals were invited to the study venue (community facilities PCYC Queensland, a not-for-profit sports and recreation organisation) to complete written informed consent and baseline assessments. To ensure allocation concealment, randomisation (block size of two generated using randomizer.org) was overseen by a researcher not involved with data collection (SP) and performed after completion of baseline assessments.

### Intervention procedure

The study groups were motivational PA counselling (MOT) or supervised gym-based exercise (GYM) interventions. The GYM and MOT interventions were facilitated by an accredited exercise physiologist (AEP) and researchers with tertiary qualifications in a health-related field (e.g., physiology, public health). Interventions were 8-weeks in duration, involving one 60-minute session/week in groups of up to 10 participants. Both interventions were designed to enhance capability, opportunity, and motivation for PA using intervention functions identified in the Behaviour Change Wheel framework (Michie, van Stralen, et al., 2011). Specific behaviour change techniques identified in the CALO-RE taxonomy (Michie, Ashford, et al., 2011) are provided in Supplementary information (Table S.1). An overview of the intervention structure is provided in Fig. 1; a brief description is provided below.

**Fig. 1:**
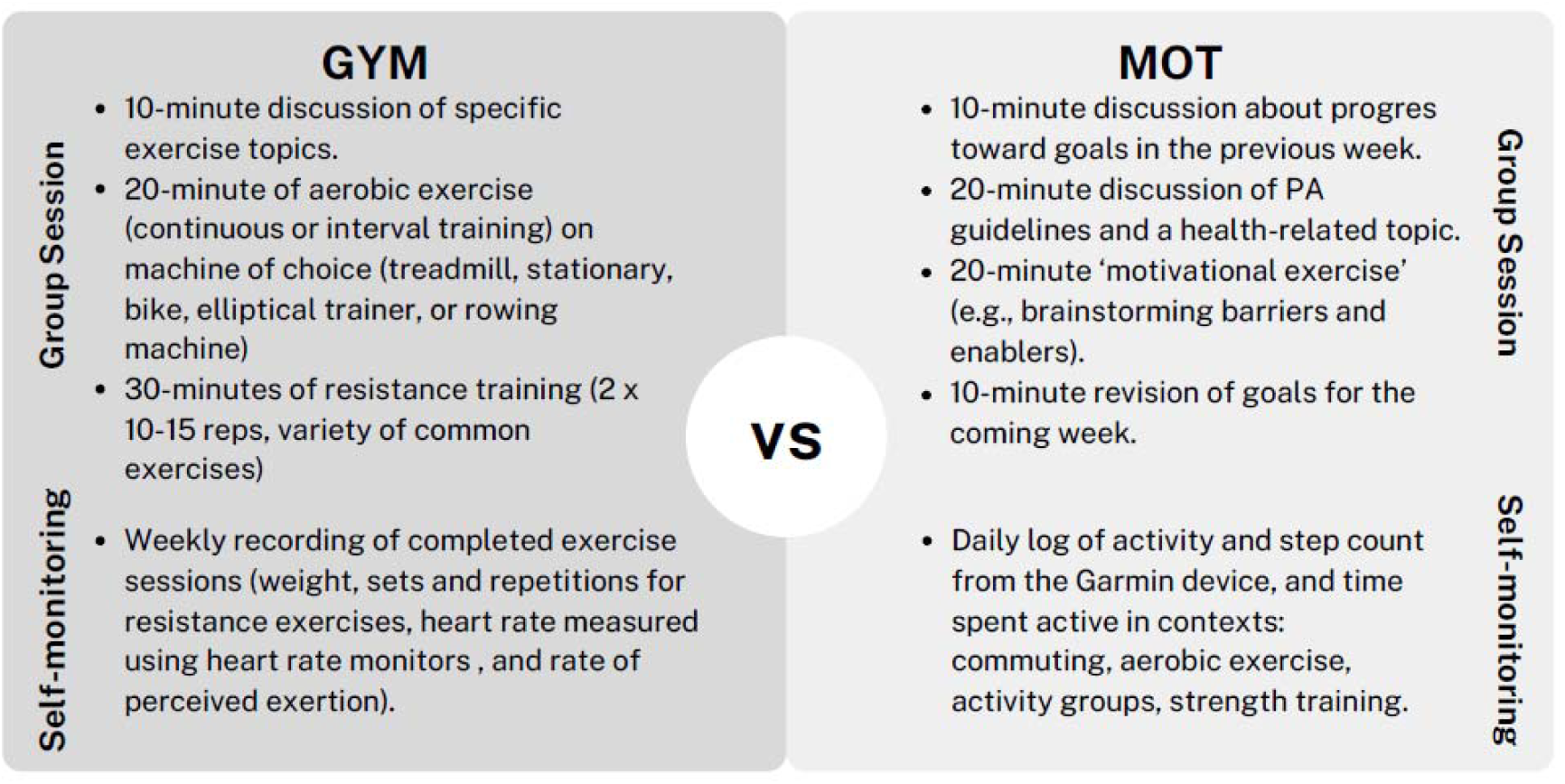
MOT and GYM intervention structure.

#### Motivational intervention (MOT)

Participants were provided Garmin Vivofit 3 devices which provide real-time feedback about daily steps, distance walked, energy expenditure, and time spent in MVPA per week. Participants were asked to keep a logbook of activity completed. Motivational sessions involved goal setting and reviewing goals each week, discussion about PA guidelines and a health-related topic, and a ‘motivational exercise’ (e.g., brainstorming barriers and enablers, positive and negative self-talk). Data from the Garmin devices or weekly logbooks were not used for analysis.

#### Gym exercise intervention (GYM)

Participants were provided gym memberships at no cost, and weekly group sessions were delivered by an accredited exercise physiologist (AEP). The intervention was based on PCYC’s ‘Healthy Bodies, Healthy Minds’ program, which progressively introduces participants to gym-based exercises (aerobic and resistance training) to promote exercise confidence in developing their exercise program based on personal abilities and preferences in consultation with the AEP. Participants were asked to record exercises completed each week in a logbook; however, data from the logbooks were not used for analysis.

### Data collection

Participants were offered gift cards to the value of $20, $30, and $40 Australian dollars for completing assessments at baseline, post-intervention, and follow-up, respectively. Post-intervention assessments were administered by a researcher blind to participant allocation. All participants were invited to complete the post-intervention assessments regardless of discontinuation with the intervention. Information about the assessments is provided below.

#### Participant characteristics

Mental health clinicians (psychiatrist or case manager) provided psychiatric diagnosis at referral (ICD-10-CM codes). Education, employment, income management, and sex were assessed using a health and demographic questionnaire. Intervention preference was also assessed by asking *‘Given what you know about the study conditions, which one would you prefer?’*.

#### Accelerometry

Bodily acceleration was measured using GENEActiv Original accelerometers (GENEActiv, Activinsights Ltd, Kimbolton, UK), which are waterproof devices requiring no user input, and similar in appearance to a wristwatch. They do not provide feedback thereby limiting the potential for reactivity (Clemes et al., 2008; Clemes et al., 2009). The sampling frequency was set at 10Hz to extend the battery life to up to 60 days. Data were analysed with R-package GGIR (https://cran.r-project.org/web/packages/GGIR/) v2.1–0 (Migueles et al., 2019). Non-wear time was defined using GGIR defaults as 60-minute periods of inactivity based on the in which the standard deviation of at least two axes is below the noise level of the accelerometer (13 mg) and the value range is lower than 50 mg; invalid data were imputed by the average at similar times of different days of the monitoring period (Van Hees et al., 2013). Light, moderate and vigorous activity were defined using thresholds of 40 mg (Hildebrand et al., 2017), 100 mg, and 400 mg (Hildebrand et al., 2014), respectively. Time spent in MVPA was calculated by summing time classified as moderate and vigorous activity. Participants were asked to wear monitors on their non-dominant wrist 24 hours/day for seven consecutive days at baseline, and continually during the 8-week intervention period. Weekly estimates for time spent in sleep, sedentary behaviour, light PA, and MVPA were calculated as the daily average for weeks with at least three valid days of accelerometer wear time, defined as at least 8 hours/day of valid waking accelerometer data and less than 25% of non-wear time (Trost et al., 2005). For consideration in analyses, participants self-reported their ‘relative activity’ at baseline compared with usual, with possible responses *less than*, *same as,* or *more than usual*.

#### Behavioural Regulation in Exercise Questionnaire (BREQ-3)

The BREQ-3 comprises 24 items to assess amotivation, and external, introjected, identified, integrated and intrinsic behavioural regulations (Wilson et al., 2006); relative Autonomy Index (RAI) was calculated as the weighted sum of these regulations (Koestner et al., 2008).

#### Kessler-6 scale (K6)

The K6 is a self-administered questionnaire with six items to assess general psychological distress experienced in the past month using a 5-point Likert scale, with scores over 15 indicating high distress (Kessler et al., 2002).

#### Simple Physical Activity Questionnaire (SIMPAQ)

SIMPAQ is a researcher-administered self-report questionnaire assessing time spent in bed, SB, walking, structured exercise, and average daily lifestyle PA (e.g. gardening, chores, work) in the previous week. Self-reported walking and exercise were summed to calculate MVPA (Rosenbaum et al., 2020).

### Data Analysis

Consistent with CONSORT guidelines, significance testing of baseline differences between group was not conducted (Moher et al., 2012). Instead, proportions and variance of participant characteristics and outcome measures at baseline were examined to assess potential group differences occurring by chance. Exploratory analyses involved comparing self-reported MVPA and SB with accelerometer-derived estimates using Pearson correlations. Within-group differences between baseline and post-intervention outcome measures were assessed using paired t-tests, and between-group differences in attendance and accelerometer data quality were assessed using independent t-tests. Statistical significance for all analyses was indicated at p-value <0.05 (two-sided).

To address the primary aim, the pre-registered outcome of cumulative change in MVPA from baseline in absolute minutes/week was analysed over the 9-week observation period. To assess the cumulative change in MVPA, the baseline value for each participant was subtracted from each weekly value, and summed over the intervention weeks. A linear mixed effects analysis with random slope was used to compare the difference between MOT and GYM groups, with intervention week, group, week*group interaction, and self-reported ‘relative activity’ level at baseline included as fixed effects. Primary analysis was conducted using an intention-to-treat sample, with missing data imputed using the group mean at baseline or Last Observation Carried Forward for subsequent weeks.

Because of contemporary developments in the field, we conducted additional compositional analyses addressing the primary aim to investigate whether composition of time spent in sleep, sedentary behaviour, light PA and MVPA changed over time, and whether any changes differed by intervention group allocation (Chastin et al., 2015). Because daily time spent in sleep, sedentary behaviour, light PA and MVPA will always collectively sum to 24-h/day, any increase in one behaviour must be offset by a decrease in other behaviours, meaning time-use is inherently co-dependent and compositional in nature. In order to overcome this, compositional data analysis techniques need to be used (Dumuid et al., 2018). For the compositional analysis, the 4-part movement-behaviour composition was first checked for the presence of zeroes, and then expressed as a set of three isometric log ratios (ilr) with the first ilr coordinate representing the pivot coordinate for MVPA reflecting the ratio of time spent in MVPA to the geometric mean of all other behaviours. Collectively, the ilr coordinates retain all relative information about time spent in the four behaviours yet can be used in standard multivariate statistical models. For inferential analysis, a multivariate response linear mixed model with random intercepts was used to investigate changes in composition (expressed as ilr coordinates) across the nine timepoints. This was achieved by stacking the three ilr responses into a single outcome variable along with a corresponding indicator variable (Snijders et al., 2011). Intervention week, group, and week*group interaction were added as fixed effects, and random intercepts for each participant were included to account for the repeated measurements on participants. The random intercepts and error for the three ilrs were allowed to flexibly covary by using an unstructured covariance matrix to account for the multivariate nature of the data. A type 3 multivariate analysis of variance (MANOVA) F-test was then conducted on the fixed effects of the model to determine if composition changed over time, by group, or if a significant group by time interaction was present. In order to aid interpretation, model-based estimated for the ilrs for each group and timepoints were back-transformed into the compositional space via the inverse ilr transformation and plotted.

To address the secondary aim, a linear mixed effects analysis with random intercept was used to assess the change in motivation (as an overall Relative Autonomy Index score: RAI), with intervention week, group, week*group interaction, and self-reported ‘relative activity’ level at baseline included as fixed effects. Analyses were conducted using R (v4.3.1, R Core team 2020, Vienna, Austria), and compositional analyses were conducted using the compositions package (van den Boogaart, et al, *Compositions: Compositional data analysis*; 2021) and the nlme package (Pinheiro et al., 2006).

## RESULTS

### Participant recruitment and allocation

A participant flow diagram is in Fig. 2. A total of 163 people were referred to the study between July 2018 and January 2020. Of these, 12 were unable to be contacted, 71 declined the invitation to participate, and nine were excluded for being too physically active (more than 300 min/week self-reported activity). Baseline assessments were completed by 64 participants prior to randomisation: 31 were allocated to GYM, and 33 were allocated to MOT. A higher proportion of participants in the MOT group were allocated their non-preferred condition than the GYM group (GYM=6, 19%; MOT=18, 55%), and a higher proportion discontinued the MOT intervention (GYM=4, 13%; MOT=10, 30%).

**Fig 2.**
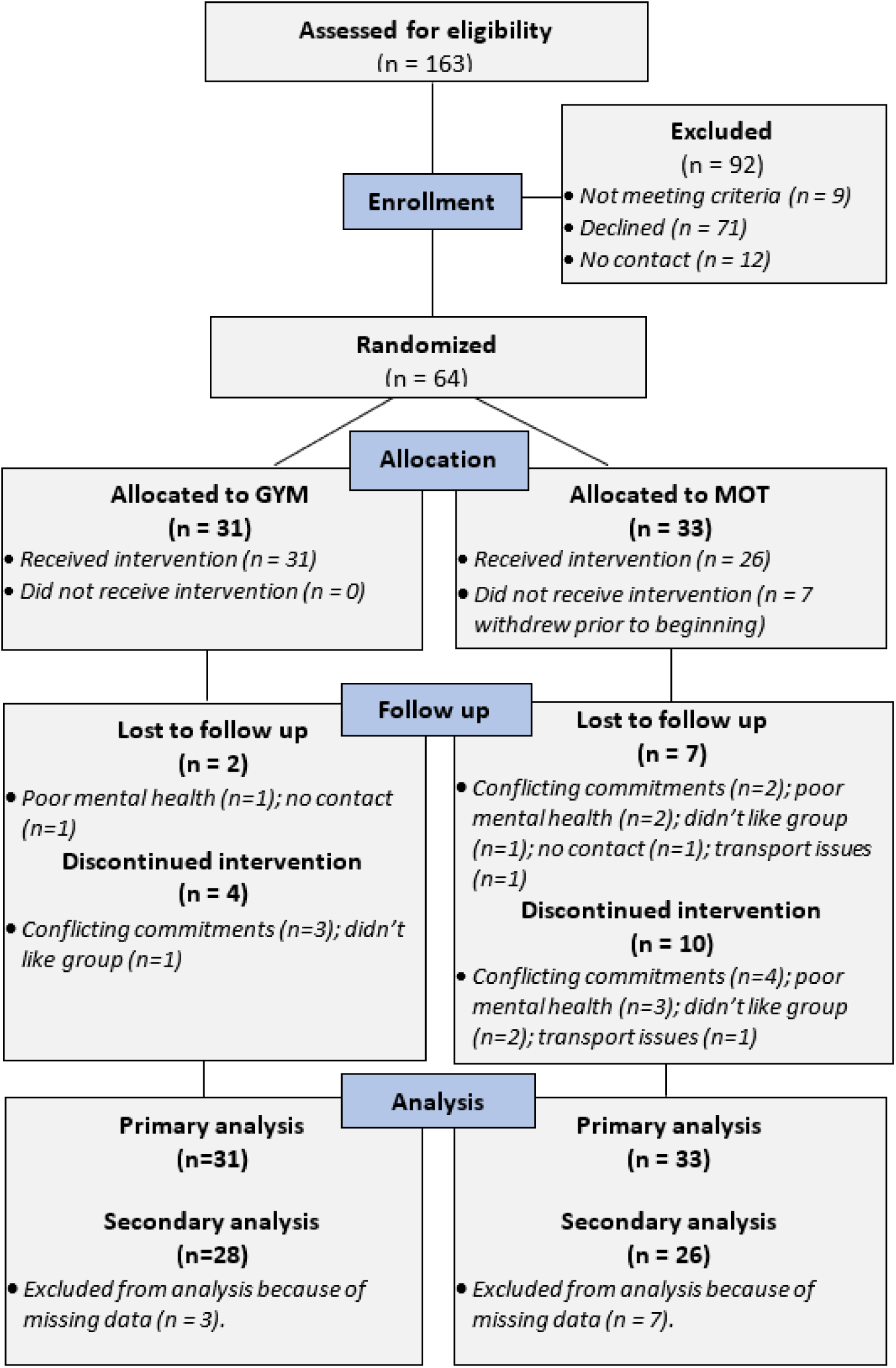
CONSORT flow diagram.

### Participant characteristics

Participant baseline characteristics are shown in Table 1. Participants had a mean age of 36 years old (range: 18 to 58 years), were predominantly male (63%), with high body mass index (BMI) (25% overweight: BMI>25kg/m^2^; 57% obese: BMI>30kg/m^2^), and roughly half (45%) had high psychological distress. Most participants had a primary diagnosis of a psychotic disorder (n=38; 59%), followed by affective disorder (n=16; 25%). Baseline characteristics were similar for GYM and MOT participants.

**Table 1:** Participant characteristics at baseline.

|  | **All (n=64)** |  | **GYM (n= 31)** |  | **MOT (n= 33)** |  |
| --- | --- | --- | --- | --- | --- | --- |
|  | *mean* | *(SD)* | *mean* | *(SD)* | *mean* | *(SD)* |
| Age (years) | 35.9 | (10.6) | 34.4 | (10.6) | 37.3 | (10.6) |
|  | range=18-58 |  | range= 18-55 |  | range= 18-58 |  |
|  | *n (%)* |  | *n (%)* |  | *n (%)* |  |
| Female | 24 | (38%) | 11 | (35%) | 13 | (39%) |
| High psychological distress | 29 | (45%) | 16 | (52%) | 13 | (39%) |
| **Number of mental health diagnoses** |  |  |  |  |  |  |
| 1 | 51 | (80%) | 25 | (81%) | 26 | (79%) |
| 2 | 9 | (14%) | 4 | (13%) | 5 | (15%) |
| 3 | 4 | (6%) | 1 | (3%) | 3 | (9%) |
| **Primary mental health diagnosis** |  |  |  |  |  |  |
| Psychotic disorder | 38 | (59%) | 17 | (55%) | 21 | (64%) |
| *with psychiatric comorbidity* | 6 | (16%) | 2 | (12%) | 4 | (19%) |
| Bipolar disorder | 11 | (17%) | 4 | (13%) | 7 | (21%) |
| *with psychiatric comorbidity* | 3 | (27%) | 0 | (0%) | 3 | (43%) |
| Personality disorders | 7 | (11%) | 4 | (13%) | 3 | (9%) |
| *with psychiatric comorbidity* | 2 | (29%) | 1 | (25%) | 1 | (33%) |
| Depressive disorder | 5 | (8%) | 4 | (13%) | 1 | (3%) |
| *with psychiatric comorbidity* | 1 | (20%) | 1 | (25%) | 0 | (0%) |
| Other (e.g., post-traumatic stress disorder; adjustment disorder) | 3 | (5%) | 0 | (0%) | 3 | (9%) |
| **Living situation** |  |  |  |  |  |  |
| Single person living alone | 24 | (38%) | 13 | (42%) | 11 | (33%) |
| Group home of unrelated adults | 11 | (17%) | 5 | (16%) | 6 | (18%) |
| Family household with only family members present | 29 | (45%) | 13 | (42%) | 16 | (48%) |
| **Education** |  |  |  |  |  |  |
| Did not complete high school | 18 | (28%) | 9 | (29%) | 9 | (27%) |
| High school | 19 | (30%) | 8 | (26%) | 11 | (33%) |
| Trade certificate/apprenticeship | 13 | (20%) | 7 | (23%) | 6 | (18%) |
| Diploma | 6 | (9%) | 3 | (10%) | 3 | (9%) |
| Tertiary degree (University) | 8 | (13%) | 4 | (13%) | 4 | (12%) |
| **Employment** |  |  |  |  |  |  |
| Unable to work | 19 | (30%) | 7 | (23%) | 12 | (36%) |
| Unemployed / looking for work | 20 | (31%) | 10 | (32%) | 10 | (30%) |
| Student | 6 | (9%) | 2 | (6%) | 4 | (12%) |
| Part-time/casual | 9 | (14%) | 6 | (19%) | 3 | (9%) |
| Volunteer | 5 | (8%) | 3 | (10%) | 2 | (6%) |
| Full-time | 1 | (2%) | 1 | (3%) | 0 | (0%) |
| Homemaker/retired | 4 | (6%) | 2 | (6%) | 2 | (6%) |
| **Income management** |  |  |  |  |  |  |
| Very difficult / Impossible | 14 | (22%) | 9 | (29%) | 5 | (15%) |
| Sometimes difficult | 20 | (31%) | 6 | (19%) | 14 | (42%) |
| Easy / 'Not too bad' | 30 | (47%) | 16 | (52%) | 14 | (42%) |
| **Smoker status** |  |  |  |  |  |  |
| Daily/occasionally | 29 | (45%) | 12 | (39%) | 17 | (52%) |
| Never/ex-smoker | 35 | (55%) | 19 | (61%) | 16 | (48%) |

| **BMI (kg/m<sup>2</sup>)<sup>d</sup>** |  |  |  |  |  |  |
| --- | --- | --- | --- | --- | --- | --- |
| <18.5 | 1 | (2%) | 1 | (3%) | 0 | (0%) |
| 18.5 – 24.9 | 10 | (16%) | 5 | (16%) | 5 | (15%) |
| 25 – 29.9 | 15 | (23%) | 4 | (13%) | 11 | (33%) |
| 30-34.9 | 17 | (27%) | 11 | (35%) | 6 | (18%) |
| >35 | 18 | (28%) | 8 | (26%) | 10 | (30%) |
| missing | 3 | (5%) | 2 | (6%) | 1 | (3%) |

(Kessler et al., 2002)

### Attendance and accelerometer data

Rates of attendance and valid accelerometer data for each week are shown in Fig. 3. Median attendance to group sessions was 4/8 (IQR=2 to 7) for GYM, and 5/8 (IQR= 2 to 7) for MOT, which was similar between the two groups t(62)=0.236, p=0.814. More than half of the group sessions (five or more out of a possible eight sessions) was attended by 13 (42%) GYM, and 19 (58%) MOT participants. The proportion of valid days of accelerometer monitoring was similar between groups t(62)=0.61, p=0.505; valid accelerometer-derived statistics were available for a median of 6 weeks (IQR=1 to 7) for GYM, and 5 weeks (IQR=1 to 8) for MOT participants. The mean average proportion of non-wear time for these valid weeks ranged from 0.9% to 2.5%. Participants’ self-reported ‘relative activity’ level at baseline was similar between groups: *less than usual* (GYM=5, 16%; MOT=7, 21%), *same as usual* (GYM=21, 68%; MOT=18, 55%), *more than usual* (GYM=5, 16%; MOT=8, 24%), χ^2^(1) = 0.135, p=0.280.

**Fig 3.**
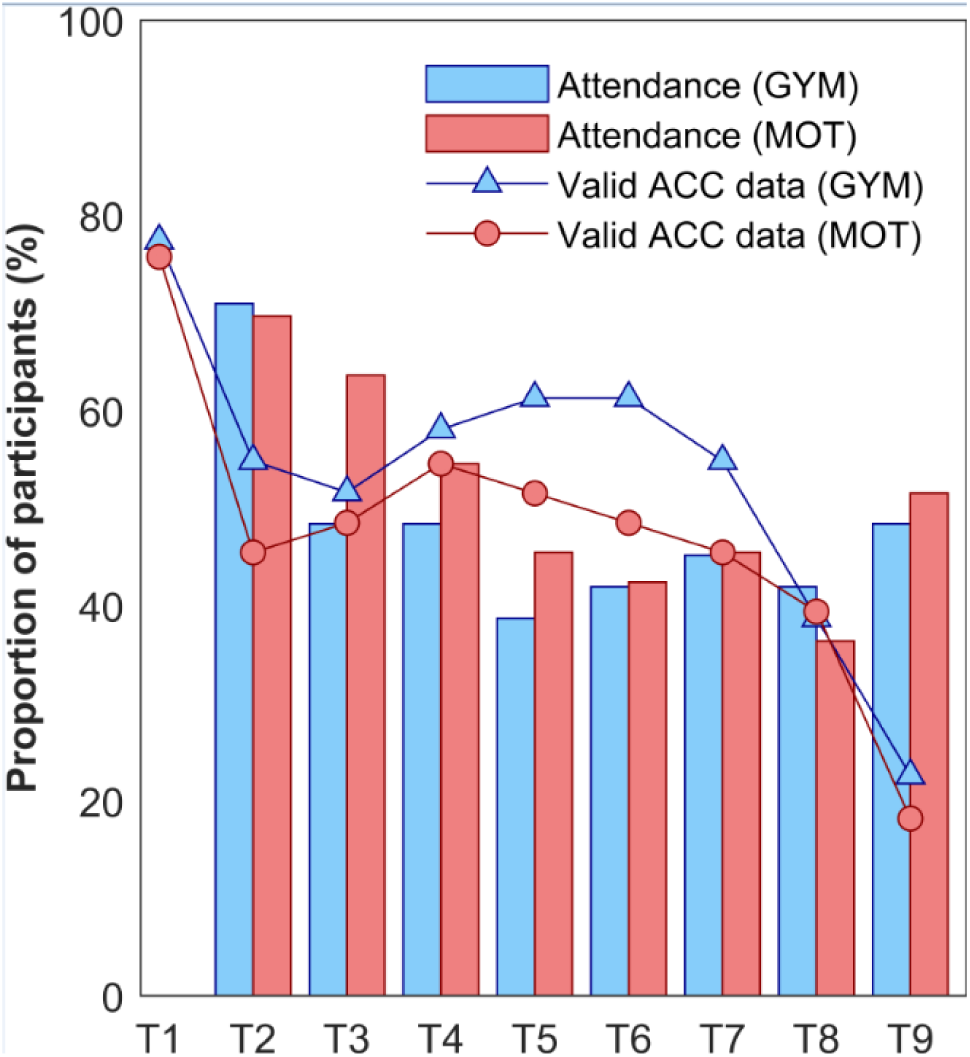
Attendance to group sessions shown as a proportion of participants allocated to GYM (n=31; blue bars) and MOT (n=33; red bars) over the 8-week intervention period (T2 to T9). The proportion of participants with valid accelerometer data (ACC) is shown for GYM (triangles) and MOT (circles) over the 9-week observation period (T1=baseline).

### Outcomes

Data for outcome measures are shown in Table 2. Considering within-group comparisons, accelerometer-derived MVPA increased for the MOT (t(32)=2.648, p=0.012) but not GYM group. Self-reported exercise increased for the GYM group (t(28)=2.047, p=0.05) and MOT group (t(25)=2.361, p=0.026), and self-reported SB also reduced for the MOT group (t(23)=2.135, p=0.044). Unadjusted mean differences (Table 2) indicated no significant difference between groups, and there were no between-group differences for any other outcome measures at post-intervention. Comparing self-report and accelerometer-derived estimates at baseline, accelerometer-derived MVPA was moderately correlated with self-reported walking (r=0.392, p=0.035) but not incidental activity (r=0.116, p=0.546) or exercise (r=-0.188, p=0.328). Accelerometer-derived SB also had moderate-to-high correlation with self-reported SB (r=0.574, p=0.001).

**Table 2:**
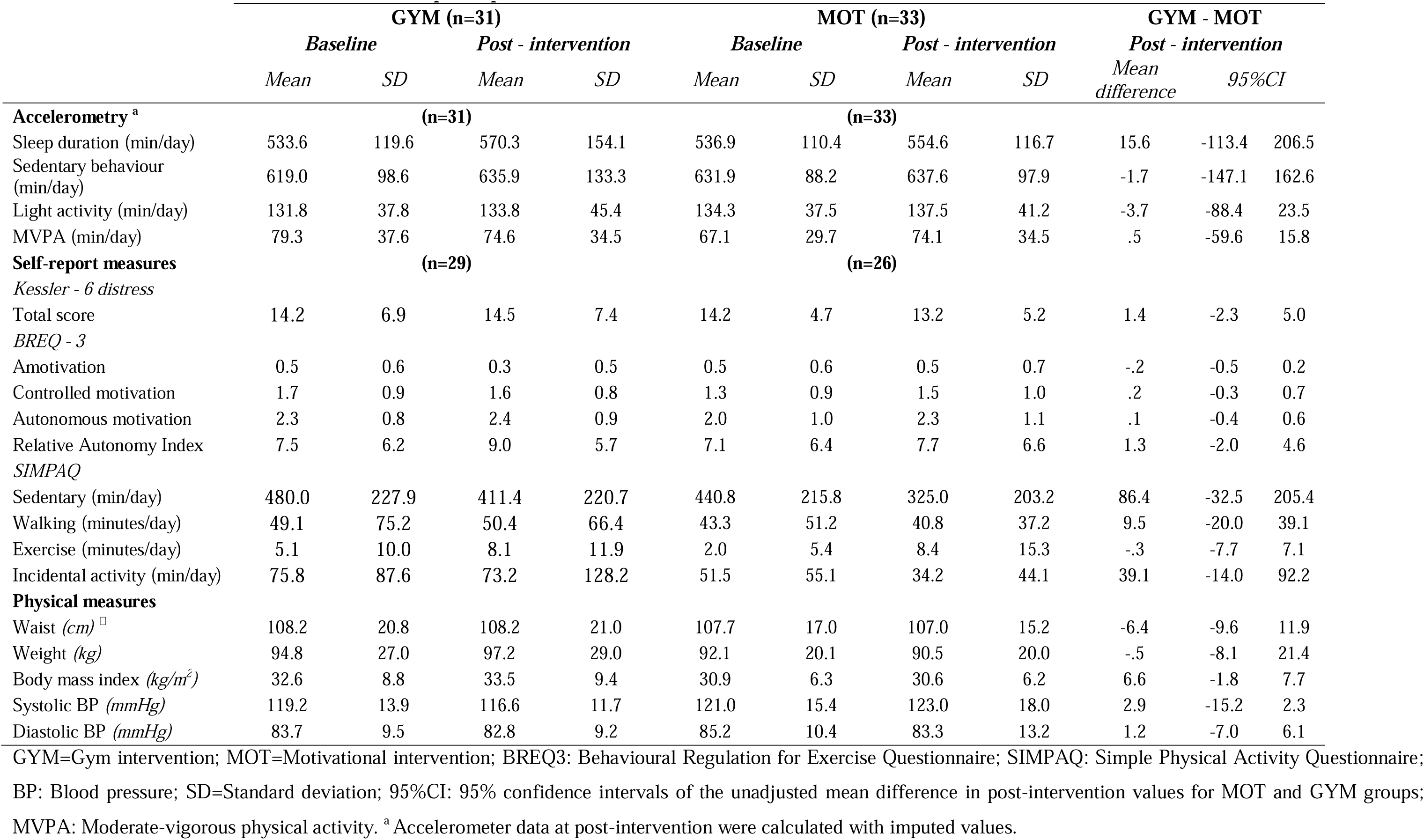
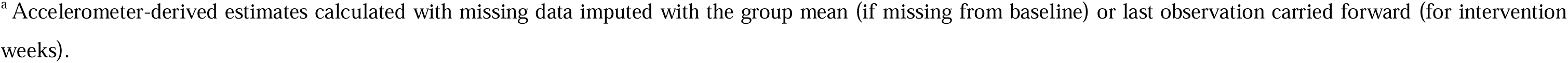
Outcome measur es for GYM and MOT participants.

### Primary analyses

Plotting the raw accelerometer-derived MVPA did not reveal any discernible trends; however, plots of the cumulative change illustrated a consistent increase in MVPA for the MOT group, with mean cumulative changes of −19.1 (SD=126.7) minutes and 49.9 (SD=131.7) minutes for the GYM and MOT groups in the ninth week, respectively (Fig. 4). The linear mixed effects model indicated a significant effect of intervention week (β=-10.86; 95%CI=−18.11 to −3.62; p=0.003), ‘relative activity’ at baseline (β =31.10; 95%CI=21.72 to 40.49; p<0.001), and week*group interaction (β =8.69; 95%CI=4.15 to 13.23; p<0.001).

**Fig 4.**
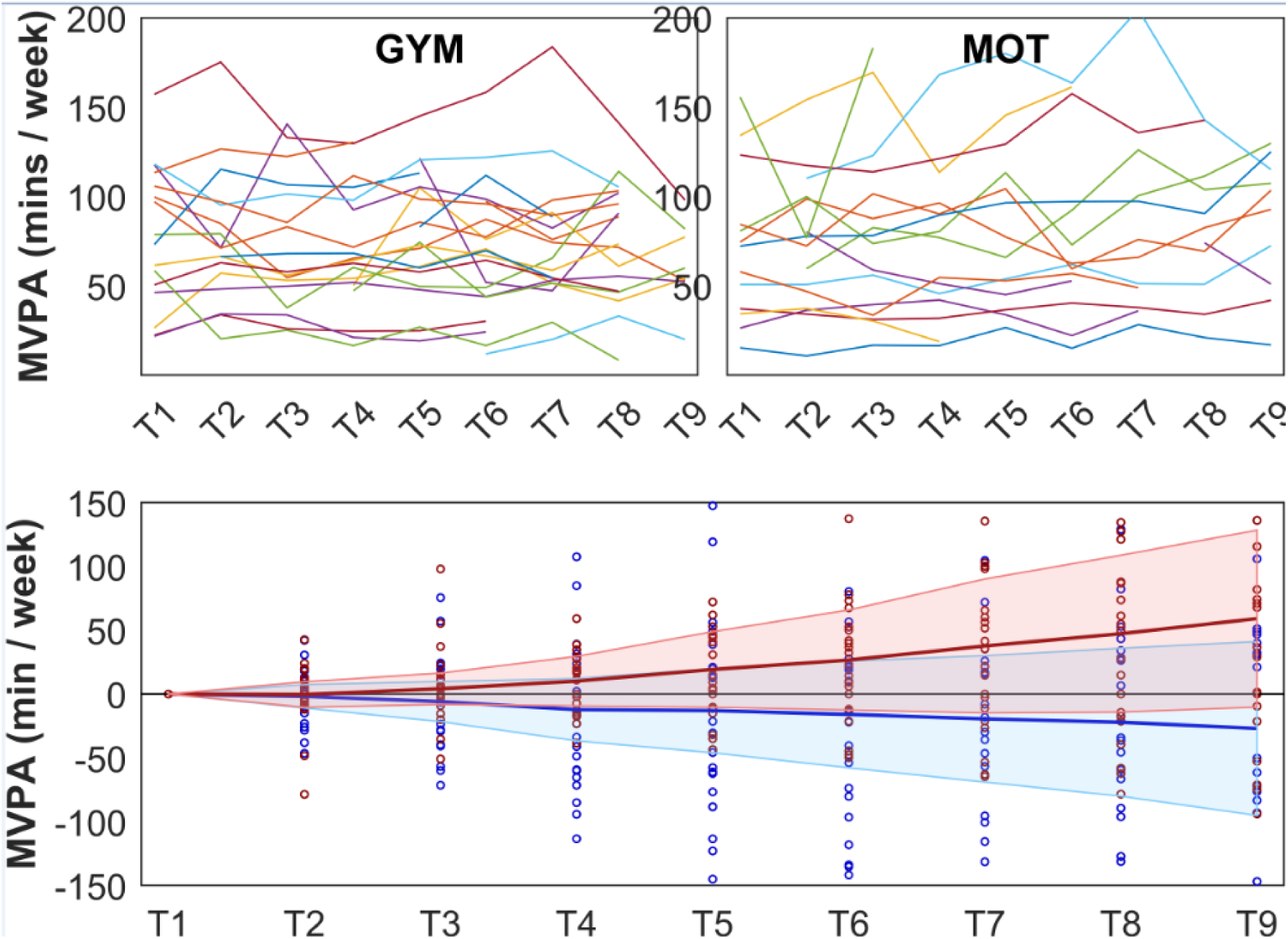
Accelerometer-derived moderate-to-vigorous physical activity (MVPA) in minutes/week plotted over the 9-week observation period (T1 to T9). Raw individual trajectories for GYM (n=31) and MOT (n=33) groups are shown in the top panels; the cumulative change for GYM (blue) and MOT (red) groups are shown in the lower panel. Cumulative change was calculated as the weekly estimates for each participant minus their baseline value (T1), hence all participants begin at zero; the central line represents the group mean and the shaded region is standard deviation.

Results for the compositional linear mixed model are shown in Table S.2. Using the Satterthwaite approximation of denominator degrees of freedom, there was no significant group by time interaction for the first ilr coordinate over the 8-week intervention, suggesting the contribution of MVPA relative to other behaviours did not differ over time by group status. Results for the MANOVA on the fixed effects of the model are shown in Table S.3. The F statistic for the group by time interaction with the vector of ilr coordinates (F(24,783)=0.6, p=0.94) suggests there was no significant difference in compositions over time by group status. Model-based estimates for time use by group are shown in Figure 5. There appear to be small increases in sleep and decreases in sedentary behaviour for both groups across the 9-week observation period; however, given results are not significant these need to be interpreted with caution.

**Fig. 5:**
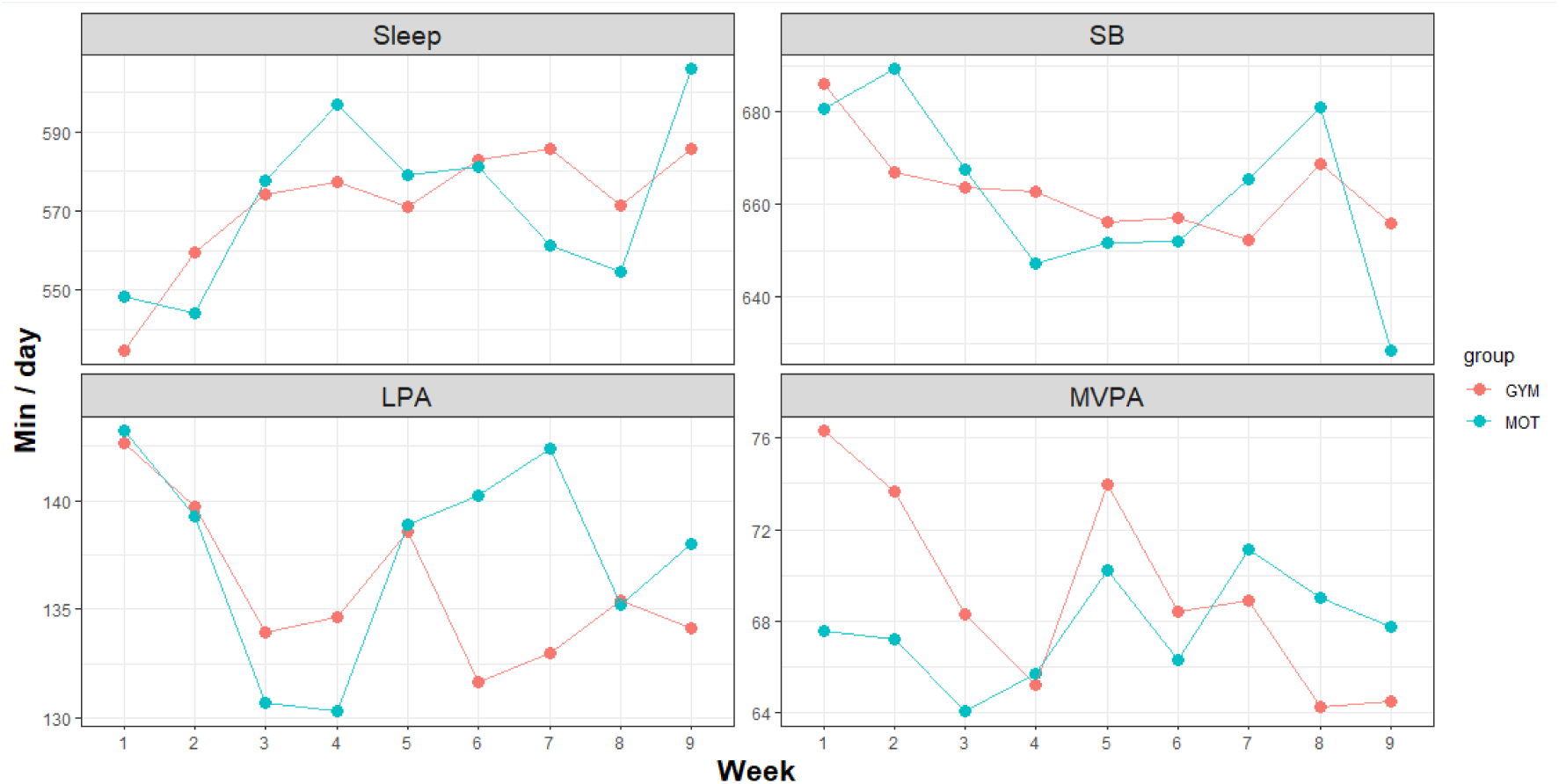
Model-based estimates for changes in activity across a the 9-week observation period for GYM and MOT groups. SB = sedentary behaviour; LPA = light physical activity; MVPA = moderate-to-vigorous physical activity.

### Secondary analyses

The mean change from baseline to post-intervention in RAI was 0.6 (SD=6.5) in the MOT group (n=26) and 1.9 (SD=6.2) in the GYM group (n=28). There were no significant effects for relative activity at baseline (β =0.48; 95%CI=-1.73 to 2.69; p=0.667), week (β =2.49; 95%CI=-2.78 to 7.77; p=0.351), group (β =-0.07; 95%CI=-5.66 to 5.51; p=0.979), or week*group interaction (β =-0.76; 95%CI=-4.12 to 2.60; p=0.656).

## DISCUSSION

MVPA is a modifiable factor related to health outcomes for people with SMI. We used innovative analytical approaches by investigating the cumulative sum of MVPA from baseline to evaluate differences in the change in total volume of MVPA during two different interventions. The cumulative change in MVPA over the intervention period was higher in the MOT group compared with the GYM group. Analyses indicated that the difference between groups was about 8.7 minutes/week, which may suggest that the motivational intervention was more effective; however, this is a marginal change overall. When inspecting within-group changes, the mean MVPA for GYM participants appeared decrease from baseline, although this was non-significant. It should also be noted that baseline MVPA as measured by accelerometry suggested that both groups were relatively active at enrolment and exceeded the 300 min/week of self-reported activity in the study protocol. Therefore, it is possible that greater improvements in MVPA would be possible with a more inactive population. To isolate potential behavioural mechanisms in this study, exercise physiologists were available to teach participants how to use the gym equipment and help them find suitable exercises; however, structured motivational strategies such as goal setting were restricted. This suggests that motivational strategies are an essential element in exercise support, and additional sources of should be incorporated in the design of exercise interventions for people with SMI (Arnautovska et al., 2022). It’s important to note that health professionals routinely employ motivational strategies within their practice; for example, a consensus statement of exercise physiology in mental health settings highlights that coaching, motivational interviewing, and goal setting are core skills of exercise physiologists (Lederman et al., 2016). Exercise physiologists can provide practical exercise support and motivational strategies to improve MVPA of people with SMI in mental health settings.

The MOT sessions were informed by the Behaviour Change Wheel and COM-B framework (Michie, van Stralen, et al., 2011), and behaviour change techniques included self-monitoring using activity trackers and self-regulatory techniques (e.g., goal setting) which have been shown to be generally effective (Brickwood et al., 2019). Goal setting was based on SMART goals in which participants were asked to set ‘performance goals’ (e.g., walk three times/week); however, the utility of this approach has been questioned for PA behaviour interventions (Swann et al., 2021). PA and exercise are complex behaviours (Swann et al., 2018) and ‘learning goals’ have been recommended when individuals are new to a complex task (Swann et al., 2021). Even though we used performance goals, we used a dynamic process by allowing participants to review goals each week and amend them on reflection (e.g., to do a different activity, or to increase or decrease the goal). This was to accommodate participants’ learning about their capabilities and preferences, and was considered essential for people with SMI who experience a variety of barriers associated with psychological distress (Chapman et al., 2016). It is possible that performance goals or the lack of established motivational strategies (e.g., motivational interviewing) in our intervention reduced effectiveness; however, there is limited evidence on motivational approaches to be used in interventions to promote MVPA in people with SMI. Of the small number that have successfully demonstrated significant behaviour change via accelerometry or pedometer measurement, all provided supervision (i.e. walking, cycling, sports such as volleyball) and not been based on PA counselling alone (Gomes et al., 2014; Ryu et al., 2020; Verhaeghe et al., 2013; Williams et al., 2019). Three of these studies offered supervision alongside weekly counselling (Verhaeghe et al., 2013; Williams et al., 2019) or incorporated behaviour change techniques into supervised sessions (Ryu et al., 2020). More studies comparing different behaviour change approaches in people with SMI will help elucidate effective intervention strategies for this group.

Compositional analyses were also presented, which provides a more complete understanding of the relative changes of MVPA in relation to light activity, SB, and sleep (Chastin & Palarea-Albaladejo, 2015). When considering the complete composition of movement behaviours (sleep, SB, light activity and MVPA), no significant differences were found between groups over time. Compositional analysis considers data that represents portions of a finite whole, i.e., time spent in MVPA is a portion of the total available time in a given measurement period, with the remaining time composed of proportions of the other behaviours (sleep, light activity, sedentary behaviour) (Chastin & Palarea-Albaladejo, 2015). This is fundamentally different from ‘conventional’ analytical methods that assume time spent in one behaviour is an absolute quantity and is essentially unbounded and independent of other behaviours. In general, the behavioural compositions of both groups were reasonably stable over time. There was no statistical evidence that any changes in behavioural compositions over the intervention period were due to group allocation, so the significant cumulative change in MVPA for the MOT group should be interpreted with caution. Our intervention parameters of 8-week duration and once/week frequency were based on our community implementation work demonstrating improvements in quality of life and self-determined motivation (Seymour et al., 2021; Whybird et al., 2020); however, this ‘dose’ of support is likely insufficient to facilitate significant behavioural changes. Previous interventions demonstrating MVPA increases have provided a greater number of sessions/week and for longer durations (Gomes et al., 2014; Williams et al., 2019) which may indicate minimally effective amounts of supervision are needed for behaviour change.

Contrary to our expectations, there was also no significant difference in motivation within- or between-groups. Our previous research comparing PA counselling and supervised exercise interventions demonstrated significant improvements in self-determined motivation, particularly for the gym group (Seymour et al., 2021). These interventions were based on the interventions trialled in the current study; however, the inclusion criteria and study design may explain differences in observed outcomes. Seymour et al. (2021) recruited participants across primary care settings and mental health services, resulting in a sample with a range of mild to severe mental health challenges. Participants of the current RCT were mostly diagnosed with psychotic disorders, and motivation may be particularly challenging for this group because of negative symptoms and associated cognitive impairments (Arnautovska et al., 2022). Further, Seymour et al. (2021) employed a quasi-experimental design in which participants were offered a choice of interventions. Self-determination theory posits that motivation can become more self-determined when psychological needs of autonomy, relatedness and competence are fulfilled, and contexts that undermine these needs thwart the internalisation of motivation (Ryan et al., 2008). The context of a clinical trial may undermine autonomy, particularly given that one-third of participants (24/64) indicated they were allocated to their non-preferred condition. There is, however, limited evidence to guide interventions designed to improve motivation for PA in people with SMI. A systematic review of 13 intervention studies reported only two studies that demonstrated improvements in motivational outcomes for people with SMI, concluding there was a lack of evidence to guide motivational interventions for people with SMI (Farholm et al., 2016).

A major strength of our study is the use of accelerometers for continual monitoring of physical movement at baseline and over the intervention period. Even though the validity of accelerometry for measuring different types of PA can vary, they can be particularly useful for assessing change in the volume of similar activity patterns over time. People with mental illness report that accelerometry is a highly acceptable form of measurement (Chapman et al., 2015) and preferred over self-report (Suetani et al., 2020). The devices were wrist-worn accelerometers which may reduce discomfort compared with devices attached to other areas such as the waist, and they had a battery life that could last up to 60 days so did not require recharging during the intervention. About three-quarters of participants provided valid data at baseline (GYM=77.4%; MOT=75.8%) which is similar to previous accelerometry research with people with mental illnesses (Chapman et al., 2015); however, only about one-fifth of participants provided valid data in the final week (GYM=22.6%; MOT=18.2%). Accelerometer data quality was a limitation: over 70% of the MVPA estimates were missing in the final week of the intervention, which reduces confidence in the findings. Accelerometers also measure physical movement at one location on the body, which may reduce the validity of comparing across different exercise types.

Compositional analysis has predominantly been used to characterise cross-sectional associations between behaviours and risk factors or health outcomes (Janssen et al., 2020; Miatke et al., 2023; Zahran et al., 2023). Fewer studies have used compositional data analysis to investigate movement-behaviour composition as a dependent variable, particularly within a repeated measures study design. Despite results of the current study suggesting no significant change in compositions over time, future studies should consider analysing movement-behaviour composition as the dependent variable, particularly in experimental and longitudinal study designs. Knowledge of how people empirically reallocate their time under different conditions will allow for more targeted intervention and public health efforts that aim to encourage favourable changes to overall composition (e.g., reallocating sedentary time towards MVPA). Finally, our dropout rate of 22% (14/64) was consistent with other exercise studies for people with schizophrenia (Vancampfort et al., 2016). Meta-analyses indicate that exercise can improve mental and physical health in people with SMI (Firth et al., 2020), and future studies should focus on the effectiveness of implementing such programs in routine care to reach a broader cohort.

## Conclusion

PA is beneficial for mental and physical health; however, it is a complex behaviour that can be challenging to adopt and maintain. While statistically significant changes in the cumulative change of MVPA were found for participants of the motivational PA counselling group, this was not confirmed with more robust compositional analyses that considers MVPA as a composition of other daily behaviours. Analysing the cumulative change of accelerometer-derived MVPA may be sensitive to small changes in behaviour; however, no change in motivation was measured in this study, suggesting the interventions delivered in a randomised trial context were not effective for longer-term behavioural changes. Motivational strategies such as goal setting and self-monitoring using activity trackers are likely more effective when combined with practical exercise support and instruction; however, more research is needed to infer optimal program characteristics (e.g. duration and frequency) and behaviour change strategies that may be effective for people with SMI. Future research should focus on combining different motivational strategies with exercise intervention and evaluating the implementation of individualised exercise programs routine mental healthcare to reach a broader cohort.

## Data Availability

All data produced in the present study are available upon reasonable request to the authors

https://researchdata.edu.au/trialling-feasibility-acceptability-mental-illness/2759877

