## Supplementary information for "A randomised controlled trial of interventions to promote adoption of physical activity in adults with severe mental illness"

1. **Intervention design**

**Supplementary Table S1: Description of intervention functions^a^ and behaviour change techniques^b^ used in the interventions**

**Gym exercise intervention (GYM)**

***Capability***

*Physical capability*

- *Training:* Verbal instruction [BCT21] and demonstration [BCT22] of different exercises by the AEP. Participants then complete the exercises with technique correction from the AEP [not coded].

***Opportunity***

*Physical opportunity*

- *Enablement*: Provision of access to exercise facility [not coded] and providing information on where and when to exercise (i.e. at the gym during opening times for supervised and unsupervised sessions) [BCT20].

*Social opportunity*

- *Environmental restructuring*: Exercise sessions delivered in groups.

***Motivation***

*Reflective*

- *Education* about purpose and general health consequences of specific exercises [BCT1], e.g. to improve posture, core stability, interval training.

*Automatic*

- *Environmental* *restructuring:* Self-monitoring of exercises completed for supervised and unsupervised sessions [BCT16].

**Motivational intervention (MOT)**

***Capability***

*Psychological capability*

- *Training:* Behavioural goal setting [BCT5] and setting graded weekly tasks [BCT9] in Week 1. Reviewing progress and reassessing goals [BCT10] each week. Refining goals in subsequent weeks, by: identifying preferred available community opportunities (e.g. activity groups; walking routes etc.) [BCT20], and action planning for PA in contexts of commuting, leisure, occupational and incidental activity [BCT7].
- *Training:* Identifying and problem-solving barriers to PA [BCT8]. Problem solving strategies include using: social support [BCT29], prompts/cures such as reminders [BCT23], environmental prompts such as getting exercise clothes ready [BCT24], use of imagery [BCT34], identifying negative self-talk and replacing with positive self-talk [BCT33], establishing routine [not coded].
- *Education:* Explanation and demonstration of strength training exercises that can be done at home [BCT21&22].
- *Education* about the processes of behaviour change [not coded], including stages of change, and internal and external motivation.

***Opportunity***

*Physical opportunity*

- *Environmental* *restructuring:* Provision of fitness tracker which provides summary feedback about activity.

*Social opportunity*

- *Environmental* *restructuring:* Exercise sessions delivered in groups.

***Motivation***

*Reflective*

- *Education* about: i) the general health consequences of PA and inactivity [BCT1], and ii) health consequences specific to adults with mental illness [BCT2], such as reducing mental illness symptoms, countering medication side-effects, and preventing physical illnesses with high prevalence in this group.

*Automatic*

- *Environmental* *restructuring:* Daily self-monitoring of PA behaviour [BCT16] using objective methods (the Garmin device) and self-report (activity log).

^a^ Nine possible intervention functions are specified in the Behaviour Change Wheel framework: *Education*, *Persuasion*, *Incentivisation*, *Coercion*, *Training*, *Restriction*, *Environmental* *restructuring*, *Modelling*, *Enablement*.[23]

^b^ Behaviour change techniques have been coded as [BCT] and numbered from the CALO-RE taxonomy;[24] techniques not listed in this taxonomy have been specified as [not coded].

2. **Linear mixed models of the compositional outcomes**

In order to model changes to the movement-behaviour composition over time by group, the 4-part composition of sleep, SB, light activity and MVPA were first expressed as a set of three isometric log ratios as below.

$$z_{1}=\sqrt{\frac{3}{4}}\ln\frac{\text{MVPA}}{\left( \text{Sleep . SB . LPA} \right)^{\text{1}\text{/}\text{3}}}$$

$$z_{2}=\sqrt{\frac{2}{3}}\ln\frac{\text{ LPA }}{\left( \text{Sleep . }\text{SB} \right)^{\text{1}\text{/}\text{2}}}$$

$$z_{3}=\sqrt{\frac{1}{2}}\ln\left( \frac{\text{SB}}{\text{Sleep }} \right)$$

Here, the first ilr coordinate reflects the dominance of time spent in MVPA relative to the geometric mean of all other behaviours. Likewise, the second coordinate represents the balance of light activity to sleep and SB, and coordinate 3 represented the balance of SB to sleep. The three ilr coordinates specified above were then used as the dependent variable of a multivariate linear mixed model with random intercepts for participants. Results for the fixed effects of the multivariate linear mixed models are shown below in Table 4. Results suggest a decrease in time spent in MVPA relative to the geometric mean of other behaviours (begative values) for the GYM group over the course of the intervention. However, these were not significant. No significant group by time interaction for any of the ilr coordinates were observed.

**Table S2. Compositional linear mixed model**

|  | Z1 | Z2 | Z3 |
| --- | --- | --- | --- |
|  | **Estimate** | **Estimate** | **Estimate** |
| Intercept | -1.37* | -1.18* | 0.018* |
| Week (vs. week 1) | -0.03 | -0.02 | -0.05 |
| Week 2 | -0.09 | -0.07 | -0.07 |
| Week 3 | -0.13* | -0.06 | -0.08 |
| Week 4 | -0.03 | -0.03 | -0.08 |
| Week 5 | -0.08 | -0.08 | -0.09* |
| Week 6 | -0.08 | -0.07 | -0.10* |
| Week 7 | -0.15 | -0.06 | -0.06 |
| Week 8 | -0.14* | -0.07 | -0.10 |
| Group (MOT) | -0.11 | -0.00 | -0.02 |
| Group*week 1 | 0.03 | -0.00 | 0.07 |
| Group*week 2 | 0.06 | -0.02 | 0.02 |
| Group*week 3 | 0.12 | -0.03 | -0.02 |
| Group*week 4 | 0.06 | 0.00 | 0.01 |
| Group*week 5 | 0.07 | 0.06 | 0.02 |
| Group*week 6 | 0.13 | 0.07 | 0.07 |
| Group*week 7 | 0.18 | 0.01 | 0.06 |
| Group*week 8 | 0.15 | 0.03 | -0.03 |

Parameter estimates for the fixed effect ilr coordinates over time and by group. * indicates significant at p <0.05

Estimates shown in Table 5. Are specific to the sequential binary partition used to construct the ilr coordinates. In order test when the whole composition of behaviours changed a multivariate analysis of variance F-test was conducted on the vector of ilrs, meaning results are invariant to the basis used when constructing the ilr coordinates. Using Satterthwaite approximation of denominator degrees of freedom, the multivariate F-test for interaction between the vector of ilr coordinates and timepoints (F(24, 783) = 0.6, p=0.94) suggests no significant difference in compositions over time by group status (Table 4).

**Table S3. Multivariate analysis of variance**

|  | Numerator DF | Denominator DF | F-value | p-value |
| --- | --- | --- | --- | --- |
| Intercept | 3 | 783 | 124.7 | <0.001 |
| Week | 24 | 783 | 0.8 | 0.80 |
| Group | 3 | 783 | 0.4 | 0.78 |
| Group*Week | 24 | 783 | 0.6 | 0.94 |

To aid interpretation the fixed effects of the model presented in Table 4. were used to estimate the vector of ilr coordinates for the two groups over the course of the nine-week intervention. The ilr coordinates were back transformed into the compositional space using the inverse ilr transformation and closed to 1440 min/day in Figure #.. Results suggest both groups showed little change in movement-behaviour compositions over the nine-week intervention period. A trend for small decreases in sedentary behaviour and small increases in sleep were observed in both groups. A small decrease in time spent in MVPA was also observed for the GYM group. But all results should be interpreted with caution given results were not statistically significant.

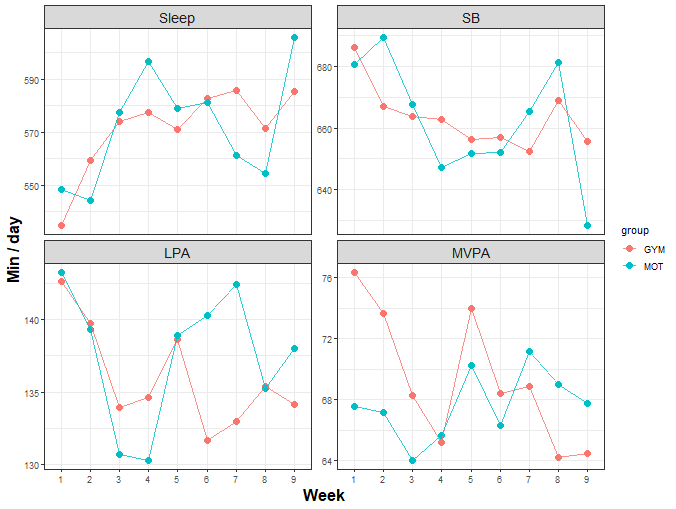

Figure #. Model-based estimates for changes in activity across a the 9-week intervention period for GYM and MOT groups. SB = sedentary behaviour; LPA = light physical activity; MVPA = moderate-to-vigorous physical activity.
